# MultiSeq-AMR: A modular amplicon-sequencing workflow for rapid detection of bloodstream infection and antimicrobial resistance markers

**DOI:** 10.1101/2024.10.25.24316117

**Authors:** Mohammad Saiful Islam Sajib, Katarina Oravcova, Kirstyn Brunker, Paul Everest, Ma Jowina H. Galarion, Manuel Fuentes, Catherine Wilson, Michael E. Murphy, Taya Forde

**Affiliations:** School of Biodiversity, One Health & Veterinary Medicine, University of Glasgow, Glasgow, United Kingdom; MRC-University of Glasgow Centre for Virus Research, Glasgow, United Kingdom; Department of Microbiology, NHS Greater Glasgow and Clyde, Glasgow Royal Infirmary, New Lister Building, Alexandra Parade, Glasgow, UK; School of Medicine, Dentistry & Nursing, College of Medical, Veterinary & Life Sciences, Wolfson Medical School Building, University of Glasgow, Glasgow, UK

**Keywords:** rapid diagnosis, amplicon sequencing, cost-effective, bloodstream infection, antimicrobial resistance, AMR, MultiSeq-AMR

## Abstract

Bloodstream infections (BSI) represent a significant global health challenge, and traditional diagnostic methods are suboptimal for timely guiding targeted antibiotic therapy. We introduce MultiSeq-AMR, an ultra-fast amplicon sequencing workflow to identify bacterial and fungal species, and a comprehensive set of antimicrobial resistance (AMR) genes (n = 91) for BSI diagnosis. We initially benchmarked MultiSeq-AMR using DNA from 21 bacterial and fungal isolates and accurately identified 100% species and 99.4% AMR genes. Further validation with 33 BACT/ALERT positive samples from suspected BSI cases revealed 100% accuracy for pathogen identification with mono-bacterial samples, with 97.4% categorical agreement (CA) for AMR gene prediction. To accelerate diagnosis, 6-hour culture-enrichment combined with MultiSeq-AMR identified 11/13 species with 96% CA for AMR gene identification. MultiSeq-AMR holds promise for improving patient outcomes as species/AMR genes could be identified in under 5 hours of BACT/ALERT positivity, and potentially <11 hours of sample collection.

## Introduction

Bloodstream infections (BSI) continue to be one of the major public health challenges worldwide, leading to high morbidity and mortality rates despite continuous surveillance efforts.^1^ The majority of these cases are traditionally attributed to Gram-positive (e.g., *Staphylococcus*, *Streptococcus* and *Enterococcus*) or Gram-negative (e.g., *Enterobacteriaceae* and *Pseudomonas*) bacterial species, however, reports of fungal and viral causes are also on the rise.^1–3^ As multidrug resistance (MDR), mainly in Gram-negative bacteria, is on the rise^1^, timely diagnosis and appropriate antimicrobial therapy are crucial for better patient outcomes in BSI, especially for patients developing early signs of sepsis.^4^ Identification and antimicrobial susceptibility testing by traditional culture-based methods, with typical time to positivity up to 48-72 hours, are not fast enough to guide targeted antibiotic therapy. Therefore, empirical antibiotic treatment plays a significant role in the early stages of clinical management.^5^ Though strict adherence to clinical guidelines can lower mortality of hospitalised critically ill patients, individuals infected with resistant pathogens are less likely to receive the appropriate antibiotic treatment initially, which may lead to poor clinical outcomes and contribute to the selection of antimicrobial resistance (AMR).^6^ With the looming rise of MDR bacterial pathogens, such cases are likely to increase over time. Therefore, rapid and sensitive diagnostic methods are urgently required to better inform antibiotic regimens and preserve the limited number of critically important antimicrobials.^6–9^

Next generation sequencing (NGS) is a promising alternative for BSI diagnosis, as certain NGS workflows can rapidly identify bacterial species and/or AMR determinants.^10,11^ However, one of the biggest challenges of NGS, specifically unbiased metagenomic next-generation sequencing (mNGS) for BSI diagnosis, is the proportion of host DNA compared to the pathogen. Host represents >99.9% of total nucleic acid in blood samples, and therefore, direct mNGS results in reduced analytical sensitivity, increased sequencing cost and time per sample to identify both pathogen and AMR determinants for rapid patient management.^12^

To improve performance, several studies have employed targeted amplicon sequencing (TAS) for detecting bacterial/fungal species and/or antimicrobial resistance genes from different types of samples, where host proportions are significantly higher than those of the pathogen.^9,13,14^ While this approach is sensitive, cost effective and hence shows promise for BSI diagnosis, one of the major limitations of TAS is its lack of comprehensiveness compared to mNGS in identifying AMR determinants. To date, most studies utilizing TAS have included a smaller collection of AMR targets common for a particular species.^14,15^ Multiplex AMR sequencing panels with more comprehensive collection of targets have been described in the literature, however, none of them have been developed/tested for diagnosing BSI in combination with rapid sequencing platforms for deployment in a clinical laboratory setup.^16,17^

This study presents MultiSeq-AMR, a rapid, cost effective, and modular Nanopore amplicon sequencing based workflow that can identify bacterial and fungal species and a large number of clinically important AMR genes from blood or other culture-enriched samples. We initially developed and benchmarked this workflow using extracted genomic DNA from reference bacterial/fungal isolates. Later this method was validated with BACT/ALERT positive and negative culture-enriched samples from individuals showing symptoms related to BSI. To accelerate microbiological diagnosis, MultiSeq-AMR was finally tested on rapid culture-enriched whole blood spiked with clinically important bacterial species.

## Results

### MultiSeq-AMR identifies pathogen and AMR determinants with high accuracy

Barcoded and cleaned PCR pools prepared from genomic DNA of pure bacterial/fungal isolates had 17.5-fold higher DNA concentration (Positive: 51.93 ng/μL versus control: 2.96 ng/μL) on average compared to human DNA and negative controls (**Fig. 1A**). All the bacterial (16/16) and fungal (5/5) isolates tested were identified accurately at both genus and species level. Both host DNA and H_2_O negative controls did not yield contigs after assembly and quality filtering and therefore identified as negative (3/3). A median of 86.66% (St. dev ± 22.24%) blast contig hits (ssciname) per sample were assigned to the correct species used for benchmarking (**Fig. 1B**). With 10 Mbp sequencing yield per sample, MultiSeq-AMR identified all 21 positive and 3 negative samples accurately. Even with only 2 Mbp yield, 95% (20/21) bacteria were accurately determined (**Fig. 1C**).

**Fig. 1:**
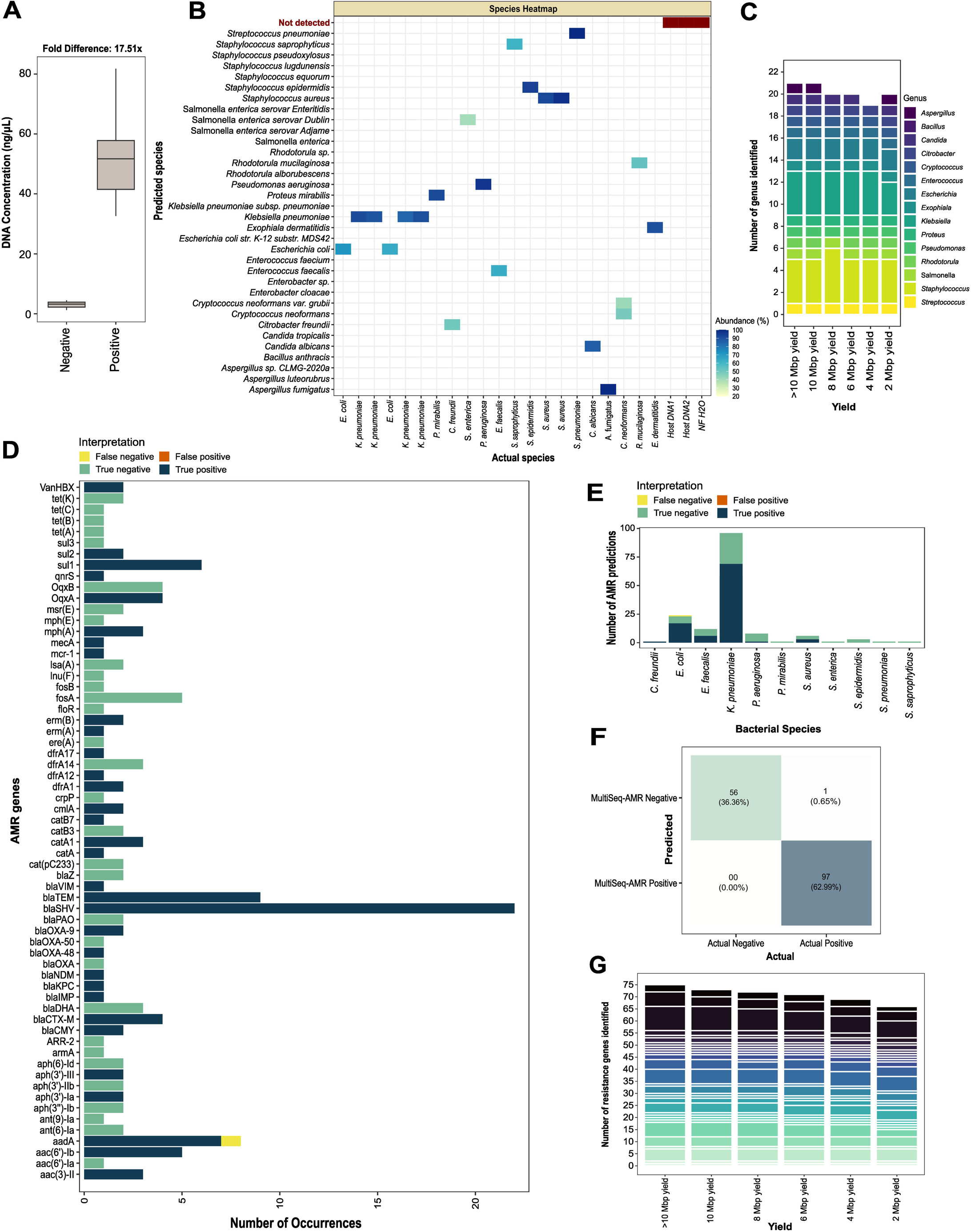
MultiSeq-AMR workflow tested on purified DNA from pure bacterial/fungal isolates. **(A)** DNA concentration of the bead-cleaned pooled PCR products, following amplification and barcoding, was measured for 21 bacterial/fungal isolates and negative controls (host DNA and H_2_O). **(B)** Bacterial/fungal species identified with MultiSeq-AMR (y-axis) versus known identity (ATCC stains) of species used for initial testing. The heatmap shows the proportion of BLAST hits matched to the species (those remaining after ≥25% abundance cutoff used in this study). **(C)** Number of genera predicted with different sequencing yields (>10 Mbp, 10Mbp, 8Mbp, 6Mbp, 4Mbp and 2 Mbp) using the same abundance cutoff. Bar plots are coloured by the type of bacterial/fungal genus identified with different sequencing yield. **(D-F)** Accuracy of MultiSeq-AMR in identifying antimicrobial resistance (AMR) genes, determined by examining four categories: false negatives (yellow: AMR primers/genes present in both MultiSeq_AMR panel and the bacteria but undetected), false positives (orange: genes/targets that are absent in the reference bacteria but detected with MultiSeq_AMR), true negatives (green: genes that should not be detected with MultiSeq-AMR, and remained undetected), true positives (deep blue: targets/genes that are present in both the panel and the bacteria and identified with MultiSeq_AMR). The same categories were compared across antibiotic genes **(D)**, bacterial species **(E)**, and combined to see overall performance **(F)**. **(G)** Number of AMR genes identified in samples with varying sequencing yield (>10 Mbp, 10Mbp, 8Mbp, 6Mbp, 4Mbp and 2 Mbp).

For AMR gene identification, MultiSeq-AMR showed 99.4% categorical agreement (153/154 predictions) overall when compared to the genes present in the reference strains. Of 98 AMR genes present among the reference strains that we expected should be predicted by MultiSeq-AMR, only one was not identified. This single false negative occurred for AMR gene *aadA* (1/8 occurrences), present in one of the *E. coli* strains benchmarked here (**Fig. 1D-F**). With 10, 8, 6, 4, and 2 Mbp yield, MultiSeq-AMR identified AMR genes with 96.05%, 94.73%, 93.42%, 90.78%, and 86.84% sensitivity, respectively, similar to the results obtained with greater (20-50 Mbp) sequencing yield (**Fig. 1G**).

### Species and AMR genescan be accuratelyidentified withBACT/ALERT positive samples

With BACT/ALERT culture media samples (n = 36), PCR amplification led to 25.4-fold difference in DNA concentration between flagged positive (n=33) and negative (n = 3) samples tested (culture positives: 47.99 ng/μL versus culture negative: 1.69 ng/μL) (**Fig. 2A**). For mono-bacterial samples, 100% (31/31) genus and 96.77% (30/31) species were accurately identified with MultiSeq-AMR. Overall, 2/4 genus and species were accurately detected from two mixed infection samples; the remaining two species identified by bacterilogical culture, i.e., *Klebsiella oxytoca* and *Enterococcus faecalis*, were not identified with our protocol. All three negative samples were called as negative, as no contigs were retained following assembly and quality filtering. A median of 81.51% (St. dev ± 21.87%) blast contig hits matched to the species identified with culture (**Fig. 2B**). With only 2 Mbp reads, bacterial prediction accuracy remained similar (>95%) to the results obtained from 20-50 Mbp sequencing yield per sample (**Fig. 2C**).

**Fig. 2:**
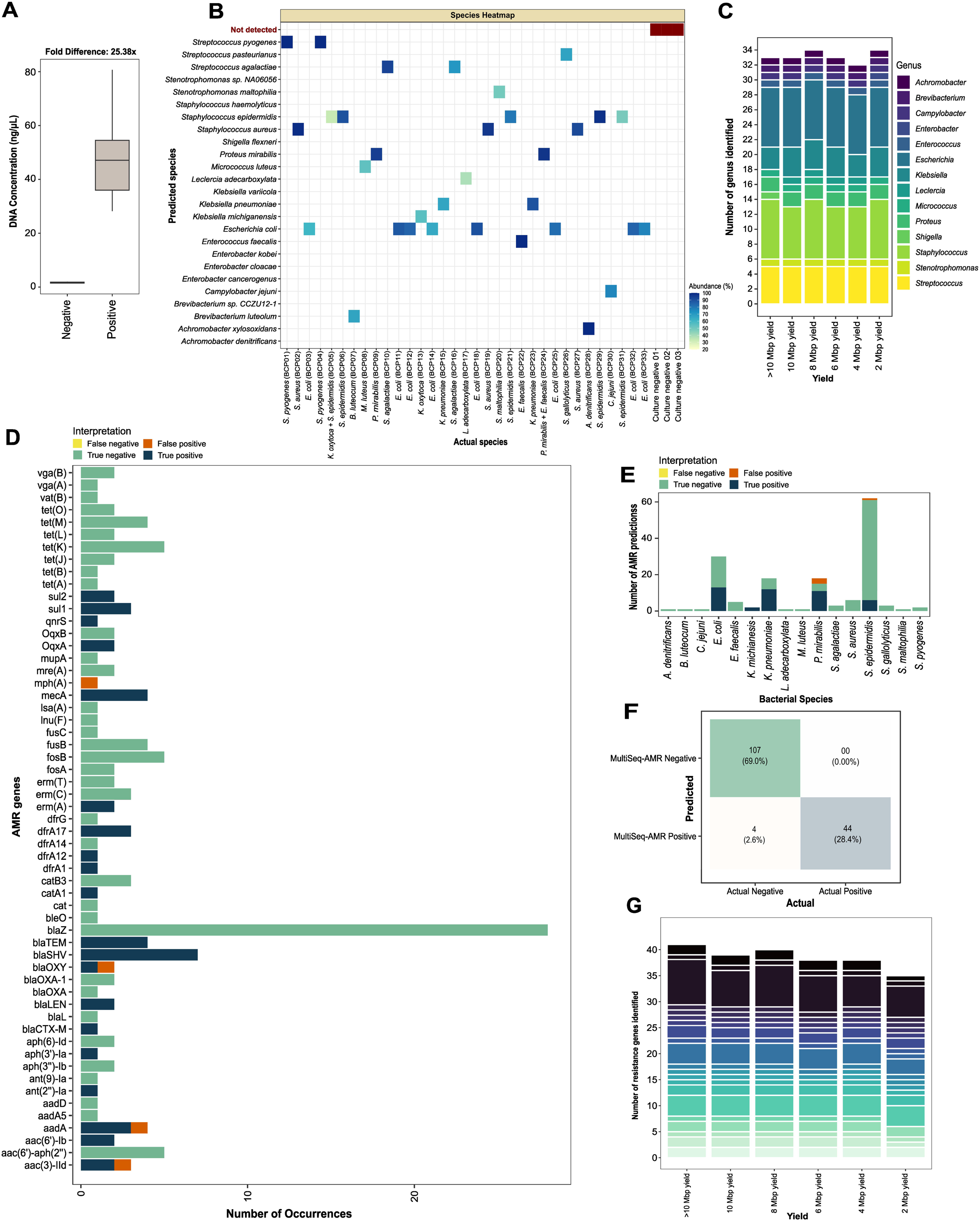
MultiSeq-AMR workflow tested on BACT/ALERT positive and negative samples. **(A)** DNA concentration of pooled PCR products, after PCR amplification and barcoding, was determined for 33 BACT/ALERT positive and three negative samples. **(B)** Pathogens identified with MultiSeq-AMR (y-axis) versus standard blood culture based results (x-axis) utilised for initial reporting. The heatmap shows the proportion of BLAST hits matched to the species (those remaining after ≥25% abundance cutoff). **(C)** Number of bacterial genera predicted with different sequencing yields (>10 Mbp, 10Mbp, 8Mbp, 6Mbp, 4Mbp and 2 Mbp) using the same threshold. Bar plots are coloured by the type of bacterial genus identified with >10 Mbp to 2 Mbp sequencing yield. **(D-F)** Accuracy of MultiSeq-AMR in identifying antimicrobial resistance (AMR) genes determined by examining four categories: false negatives (yellow: AMR primers/genes present in both MultiSeq_AMR panel and the bacteria but undetected), false positives (orange: genes/targets that are absent in the reference bacteria but detected with MultiSeq_AMR), true negatives (green: genes that are supposed to be undetected, and not identified with MultiSeq-AMR), true positives (deep blue: targets/genes that are present in both the panel and the bacteria and identified with MultiSeq_AMR). The same categories were compared across antibiotic genes **(D)**, bacterial species **(E)**, and combined to see overall performance **(F)**. **(G)** Number of AMR genes identified in samples with varying sequencing yield (>10 Mbp, 10Mbp, 8Mbp, 6Mbp, 4Mbp and 2 Mbp).

MultiSeq-AMR exhibited 97.4% categorical agreement (151/155 predictions) for predicting AMR genes in the 33 BACT/ALERT positive samples. Unlike for the reference isolates tested, no false negative predictions were observed However, resistance genes were inaccurately identified in four instances, leading to 8% (4/48) false positivity rate. Genes *mph(A)* (1/1), *blaOXY* (1/2), *aadA* (1/4), and *aac(3)-IId* (1/3) were falsely identified in two samples positive for *Proteus mirabilis* and *Streptococcus epidermidis* (**Fig. 2D-F**). Approximately 95% AMR genes (St. dev ± 2.2%) identified with 20-50 Mbp sequencing yield were still detected with 4 to 10 Mbp, and 85.36% with only 2 Mbp sequencing yield per sample (**Fig. 2G**).

### MultiSeq-AMR with rapid culture enrichmentcan facilitate same daymicrobiological diagnosis

With spiked sheep blood as host matrix, generation time for both Gram-positive (n = 7) and Gram-negative (n = 8) organisms tested for rapid culture enrichment was 22.86 (range 14.46 to 32.88 min) and 20.48 (range 16.26 to 26.04 min) minutes on average, respectively. Four-hour BACTEC enrichment led to a median concentration of 1.2×10^1^ CFU/mL for Gram-positive and 1.2×10^2^ CFU/mL in case of Gram-negative bacterial species. With six-hour culture, Gram-positive organisms reached a median of 7.0×10^2^ (from 1.15×10^2^ to 3.02×10^4^) CFU/mL and negatives 3.4×10^3^ (between 5.83×10^1^ and 2.11×10^4^) CFU/mL. Of all the species tested, *Pseudomonas aeruginosa* exhibited the slowest growth, reaching only 5.83×10^1^ CFU/mL with six-hour enrichment. Median bacterial concentration reached 2.65×10^6^ CFU/mL (between 7.50×10^4^ and 9.27×10^9^) for Gram-positive, and 9.56×10^6^ CFU/mL (from 4.33×10^5^ to 1.50×10^9^) for Gram-negative organisms with 10-hour BACTEC enrichment (**supplement Rapid_enrichment_S3**).

With six-hour rapid enrichment, DNA concentration between positive (spiked samples; n=13) and negative (sterile enriched sheep blood; n=2) samples differed 4.7-fold (positives: 20.76 ng/μL and negatives: 4.4 ng/μL) on average after PCR amplification with MultiSeq-AMR primers. Two samples, spiked with *P. aeruginosa* and *Streptococcus pneumoniae*, had significantly lower DNA concentration (average 8.95 ng/μL) after PCR compared to other bacterial species (average 23.05 ng/μL) (**Fig. 3A**). MultiSeq-AMR combined with rapid culture enrichment identified 84.61% (11/13) bacteria accurately at both genus/species level, with a median 90.0% (St. dev ± 24.77%) BLASTn hits assigned to the species used for spiking. *P. aeruginosa* and *S. pneumoniae*, the two species yielding suboptimal amplicons/DNA concentration during PCR, were not detected, as assembled contigs did not pass the initial QC threshold for BLASTn. The two negative controls did not flag positive, as expected (**Fig. 3B**). The accuracy of bacterial species identification was the same with as low as 4 Mbp yield as obtained with 20-50 Mbp yield per sample (84.61%; 11/13 species). With 2 Mbp sequencing yield, MultiSeq-AMR detected 10/13 bacteria accurately. *Enterococcus faecium*, in addition to *P. aeruginosa* and *S. pneumoniae*, was missed (**Fig. 3C**).

**Fig. 3:**
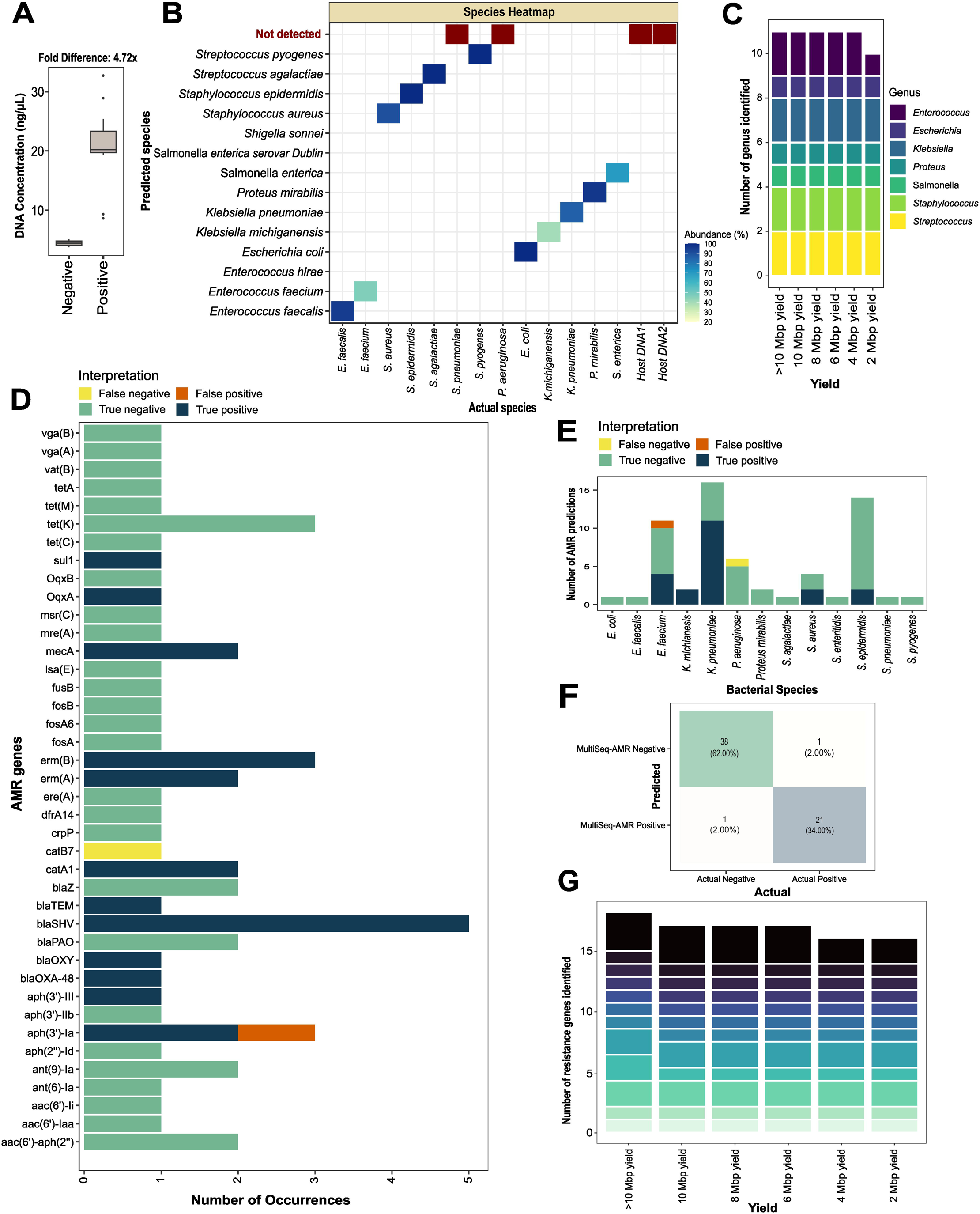
MultiSeq-AMR workflow tested on rapid culture-enriched (6-hours) spiked blood samples. **(A)** DNA concentration of pooled PCR products, after PCR amplification and barcoding, was determined for 7 Gram-positive, 6 Gram-negative bacterial species and two sterile rapid enriched whole blood samples. **(B)** Bacterial species identified with MultiSeq-AMR (y-axis) versus species initially used for spiking and rapid enrichment (x-axis). The heatmap is showing the proportion of BLAST hits matched to the species remaining after ≥25% abundance threshold. **(C)** Number of genera predicted with different sequencing yields (>10 Mbp, 10Mbp, 8Mbp, 6Mbp, 4Mbp and 2 Mbp) using the same threshold. Bar plots are coloured by the type of bacterial genus identified with varying sequencing yield (>10 Mbp to 2 Mbp). **(D-F)** Accuracy of MultiSeq-AMR in identifying antimicrobial resistance (AMR) genes determined by examining four categories: false negatives (yellow: AMR primers/genes present in both MultiSeq_AMR panel and the bacteria but unidentified), false positives (orange: genes/targets that are absent in the reference bacteria but detected with MultiSeq_AMR), true negatives (green: genes that are not supposed to be detected by the panel, and remained undetected), true positives (deep blue: targets/genes that are present in both the panel and the bacteria and identified with MultiSeq_AMR). Same categories were compared across **(D)** bacterial species, **(E)** antibiotic genes, and **(F)** combined to see overall performance. **(G)** Number of AMR genes identified in samples with different sequencing yield (>10 Mbp, 10Mbp, 8Mbp, 6Mbp, 4Mbp and 2 Mbp).

Overall, 96% categorical agreement (59/61 predictions) was observed for AMR gene identification. One false negative (1/39), and one false positive (1/22) prediction were obtained from genes *catB7* and *aph(3”)-Ia* respectively. *E. faecium and P. aeruginosa* were two of the bacterial species exhibiting false positive and false negative gene predictions (**Fig. 3D-F**). With 10, 8, and 6 Mbp yield, 94.1% (16/17) genes matched with the 20-50 Mbp yield. Up to 88.2% concordance was seen with only 4 and 2 Mbp sequencing yield (**Fig. 3G**).

## Discussion

Bloodstream infections are among the leading causes of mortality and morbidity worldwide and require timely microbiological diagnosis for more precise and effective antibiotic treatment leading to better patient outcomes. In working towards that aim, this study introduced MultiSeq-AMR, a rapid nanopore amplicon sequencing workflow to detect bacterial and fungal species and a comprehensive set of antimicrobial resistance determinants to aid and accelerate existing methods for diagnosing BSI. MultiSeq-AMR leverages the rapid, high throughput and real-time sequencing capability of Oxford Nanopore Technologies platforms. The use of rapid PCR barcoding and library preparation steps enables sample preparation in approximately 4 hours, with sequence data available for analysis and possible result reporting almost immediately after sequencing begins. The choice of oligonucleotides includes two universal bacterial (16S rRNA) and fungal (28S rRNA), and 91 clinically important AMR gene targets, covering 11 antibiotic classes. We validated MultiSeq-AMR using reference fungal and bacterial strains, BACT/ALERT positive and negative clinical samples from individuals with suspected BSI, and rapid culture enriched spiked whole blood samples.

There are various bioinformatic options available for identifying bacterial and fungal species utilizing 16S/28S rRNA reference datasets.^18,19^ However, only a few have been standardised for BSI diagnosis, and/or tailored to work on mixed amplicon sequencing reads. Therefore, benchmarking these workflows was not within the scope of this study. Instead, we utilised a simple bioinformatic approach that utilises some of the well reputed tools to identify species and AMR determinants to evaluate the utility of MultiSeq-AMR. Later this workflow/cutoff can be modified or replaced with newer alternatives if required. Of all the possible cutoffs, ≥25% abundance threshold of BLAST contig hits showed the highest sensitivity and specificity for genus and species detection (data not shown). With this threshold, MultiSeq-AMR detected 21 bacterial/fungal reference strains, and 31 mono-bacterial BSI samples with a genus and species level accuracy of 100% and 98.4%. Even with as little as 2 Mbp sequencing yield per sample, genus level prediction was ≥95%. For two BACT/ALERT mixed species samples (BCP05, and BCP24) having two bacterial species each, only one was detected. This is likely due to the low abundance of the two species that were missed (*Klebsiella oxytoca* and *Enterococcus faecalis*) compared to the other two (*Streptococcus epidermidis* and *Proteus mirabilis*) identified with MultiSeq-AMR. With a reduced abundance threshold, we were able to recover both *K. oxytoca* and *S. epidermidis* from sample BCP05, however, this adjustment also resulted in reduced specificity in other samples (data not sown). As a result, it was decided to keep the existing cutoff (≥25%) to achieve the best overall specificity. For the rapid (6-hour) culture-enriched spiked blood, species from 11 of 13 positive samples were correctly identified (sensitivity: 84.61%). No bacteria were detected from two samples spiked with *Pseudomonas aeruginosa* and *Streptococcus pneumoniae*, exhibiting false negativity. This may be a result of slow growth rate of some of the species/stains used for the rapid enrichment experiment. For example, P. *aeruginosa* reached 5.83×10¹ CFU/mL after six hours of BACTEC enrichment. This concentration is possibly too low for the DNA extraction and PCR kit to reliably extract/amplify target regions. These two species also had significantly lower DNA concentration after PCR barcoding (8.95 ng/μL versus 23.05 ng/μL for other samples). This means the 6-hour enrichment method may not sufficiently enrich slow growing bacterial/fungal species^20^, and therefore be falsely identified as negative. However, because MultiSeq-AMR is aimed to be used alongside traditional methods, it is likely that these missed organisms would be identified later with culture. It is also worth noting that there might also be many bacterial species not tested with the rapid enrichment method that could potentially reach the desired threshold for robust capture and detection with MultiSeq-AMR. A large collection of clinical samples with multiple strains/genotypes from the same species would ideally be used to more fully validate rapid enrichment with MultiSeq-AMR in the future.

Similarly to the approaches described for species identification, there are multiple methods available for detecting AMR genes from sequencing reads.^21–23^ Again, rather than assess multiple tools, we implemented a well reputed tool/database, ResFinder, to preliminarily test the utility of MultiSeq-AMR. While comprehensive benchmarking could help identify which bioinformatic tool has the best overall performance, this was outside the scope of the present study. By default, ResFinder 4.0 has a “Threshold for %ID” of 90%, and 60% “Minimum length” cutoff. Because MultiSeq-AMR generates smaller amplicons that could fall well below the default threshold for some targets (e.g., *MecA, MecC*), we gradually reduced these values to determine those that achieved the greatest accuracy, which resulted in the selection of 40% “Threshold for %ID” and “Minimum length”. With this selected cutoff, MultiSeq-AMR achieved 98.4% categorical agreement overall for extracted genomic DNA and BACT/ALERT positive samples across 49 AMR targets, grouped into seven primer pools. Even with 6-hour rapid culture enrichment, 59/61 expected AMR genes were correctly identified, with only one false negative and one false positive prediction. Up to 85% genes were identified with only 2 Mbp sequencing yield for all the sample types, meaning the sequencing time to generate AMR and species reports could be almost instantaneous following a sequencing run.

A somewhat costly disadvantage for the rapid enrichment method could be the requirement to sequence all enriched blood samples from suspected BSI patients blindly until sufficient sequencing reads are generated for analysis in order to determine sample positivity or negativity (since these would be tested before cultures bottles flag positive). However, when we compared positive (containing pathogen) and negative samples (culture-negative or sterile blood sample), the DNA concentration after PCR barcoding and cleanup was at least 5 to 20 times higher in samples containing detectable DNA quantities of a given pathogen. This provides a unique signature which could potentially be exploited to select and sequence only the samples with significantly higher post-PCR DNA concentration compared to the negative controls to save costs. However, this approach must be validated/calibrated site-wise using more patient samples, including both positive and negative cases, across different clinical contexts before implementation.

It is important to acknowledge a few limitations of this study. Of the 13 AMR pools designed as part of this study, we successfully benchmarked seven (n = 49 targets) using reference fungal and bacterial strains, BACT/ALERT positive and negative samples, and rapid (6-hour) culture-enriched spiked samples. The remaining six AMR pools covering 42 gene targets could not be validated due to the lack of sufficient positive controls. In future, the validation of these additional pools (8 to 13) with appropriate positive and negative controls would greatly expand the AMR gene panel that could be tested with this approach. Because all the primers/pools were designed similarly, it is likely that most primer sets would amplify successfully, but primers failure or reduced amplification efficiency cannot be ruled out. Also, the MultiSeq-AMR workflow was validated using R9.4.1 flow cells and supported sequencing kits, whereas more accurate R10.4.1 kits became available halfway through the study. We believe MultiSeq-AMR can easily be adapted for R10.4.1 flow cells and kit chemistries and the overall performance would benefit from more accurate (≥Q20) sequencing reads for predicting species and AMR determinants, although this must be validated and confirmed. Taken together, this study demonstrated that MultiSeq-AMR is highly effective in detecting bacterial and fungal species, and clinically important antibiotic-resistant genes from samples such as extracted genomic DNA, BACT/ALERT flagged positive culture media, and rapid culture-enriched samples for a large group of clinically relevant pathogens. With only about 4- hours of hands-on time, MultiSeq-AMR can enable microbiological diagnosis under five hours of BACT/ALERT positivity or 11 hours of sample collection, given the availability of a standardised real-time bioinformatics workflows. The choice of AMR targets can be tailored and extended indefinitely, pathogen/syndrome-wise, and hands-on time could be further reduced through automation, making this approach highly modular. Consequently, as a rapid, cost-effective, and modular workflow, MultiSeq-AMR has potential in improving outcomes for critically ill patients with BSI.

## Methods

### Reference strains, substitute host matrixand patient samples

Sterile whole blood from healthy human volunteers and sheep (E&O Laboratories, Scotland, United Kingdom) collected in tubes containing EDTA were used as initial substitute for infected blood samples to benchmark MultiSeq-AMR. The list of clinical isolates and ATCC strains used in this study for spiking, enrichment and other experiments can be found in **Supplement Sample_info_S1.** Blood culture positive (n =33) and negative (n = 3) samples from individuals suspected of having BSI were surplus specimens collected initially as a part of routine diagnostic testing.

### Primer design to identify species and antimicrobial resistance genes

Current version of MultiSeq-AMR is comprised of 93 targets in a total of 14 primer pools: one pool with two amplicon targets to identify bacterial or fungal species, and 13 AMR pools, each targeting up to 7 unique genes per pool (**Fig. 4**, **Supplementprotocol: MultiSeq_AMR**). The species specific pool contains primers to amplify 16S rRNA^24^ and 28S rRNA^25^ for bacterial and fungal species identification, respectively. AMR pools include 91 unique gene primers from 11 antibiotic classes (**Supplement Primer_Seq_S2**).

**Fig. 4:**
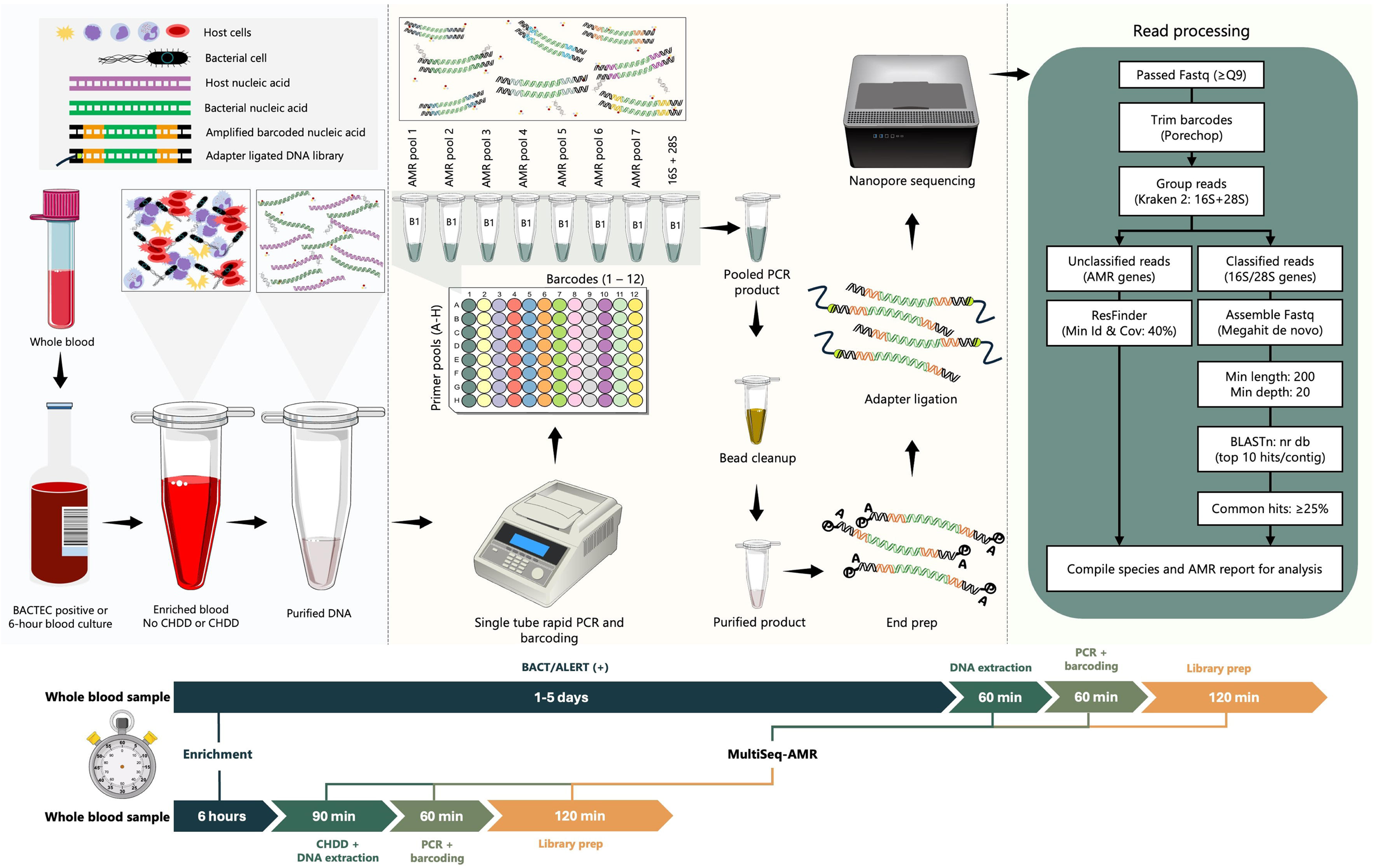
Schematic diagram highlighting major steps in MultiSeq-AMR workflow including DNA extraction, PCR barcoding, library preparation and sequencing and data analysis. Both BACTEC positive and rapid enriched (6-hour) blood culture samples can be used, each with slightly different initial processing steps. BACTEC positive samples can be subjected to DNA extraction directly, however, rapid culture-enriched samples must undergo chemical host DNA depletion (CHDD) to reduce host DNA and concentrate bacteria in the sample prior to nucleic acid purification. Total hands-on time for MultiSeq-AMR is 4 to 4.5 hours for 6-8 samples including CHDD and/or DNA extraction, PCR barcoding, and library preparation. The PCR plate example is showing the arrangement of the seven AMR and one species-specific primer pools (A-H) combined with different barcodes (B1 to B12). The first column in the plate (dark teal) highlights barcode 1 (B1).

To design MultiSeq-AMR, nucleotide sequences of all the AMR genes and variants were initially obtained from ResFinder database (bitbucket.org/genomicepidemiology/resfinder_db) (Last accessed: 30/07/2024). Each type of AMR gene was aligned using MUSCLE^26^ to identify closely related variants and differentiate between groups (e.g., *blaOXA-1*, *blaOXA-9*, *blaOXA-24*, blaOXA-48 etc) for designing primers.^26^ Subsequently, NCBI Primer-BLAST software was utilised to design AMR primers with the following parameters: a maximum primer melting temperature (Tm) of 60 °C, no cross reaction with *Homo sapiens*, and a PCR product length between 500-800 base pairs (bp).^27^ To differentiate AMR variants with sequencing, variant specific mutations were kept in the region between the forward and reverse primers for as many AMR targets/variants as possible. For genes/groups of genes where no conserved regions were found using direct Primer-BLAST, forward/reverse primer sequences were manually developed by incorporating degenerate nucleotide sequences. A total of 82 AMR primers were developed in this study and the remaining 11 primers (9 AMR targets and two targets for species identification) were sourced from previously published studies.^28–31^ The 93 primers targeting universal species/AMR genes were then tagged with outer primers having complementary sequences (5’- 3’, Forward: TTTCTGTTGGTGCTGATATTGC, Reverse: ACTTGCCTGTCGCTCTATCTTC) for PCR Barcoding Expansion 96 kit (Oxford Nanopore Technologies, Oxford, UK) and sourced from IDT (Integrated DNA Technologies, Leuven, Belgium) (**Supplement Primer_Seq_S2.)** During PCR, if multiple targets are amplified simultaneously in one pool and one AMR target is being enriched in another, this might create a significant difference in molarity per amplicon during sequencing. Therefore, to avoid significant amplicon bias, up to seven AMR targets were incorporated in each pool, ensuring that no more than three targets are amplified at once in any given pool during PCR (**Supplementprotocol: MultiSeq_AMR**). This was achieved by combining primers for gene/variants of gene that are unlikely/less likely to be present in a bacterium at the same time.

### MultiSeq-AMR workflow tested on reference bacterli/afungal isolates

A total of 16 bacterial and 5 fungal reference/ATCC strains with available whole genome sequencing (WGS) data were chosen for initial benchmarking AMR primer pools one to seven and the species-specific pool (**Fig. 4**, **Supplement Sample_info_S1**). Pure bacterial/fungal isolates were initially grown overnight in Brain Heart Infusion and Sabouraud dextrose media (Sigma-Aldrich, MO, USA). One mL overnight cultures were then centrifuged at 10,000 x *g* for 5 minutes, and the pellets were resuspended in 400 μL DNA/RNA Shield (Zymo research, CA, USA). DNA extraction was performed using QIAamp UCP Pathogen Mini kit with Pathogen Lysis Tubes L (Qiagen, Hilden, Germany) following manufacturer’s recommendation. Two sterile human blood, and one sterile H_2_O sample were also processed as negative controls alongside the 21 bacterial/fungal species. One µL extracted DNA was quantified using Qubit dsDNA broad range assay kit (Thermo Fisher Scientific, MA, USA) and 10 ng DNA were used as input/pool for PCR. Details for rapid PCR amplification/barcoding, library preparation and sequencing can be found in **Supplementprotocol: MultiSeq_AMR**.

### Validation of MultiSeq-AMR with BACT/ALERT positive samples

To further validate MultiSeq-AMR, 33 BACT/ALERT (bioMérieux, Marcy-l’Étoile, France) flagged positive blood culture (FPBC) and three negative blood culture media were processed as negative controls. All the culture positive samples had sequencing results available through a previous study for comparison with MultiSeq-AMR.^32^ Briefly, 400 μL enriched culture media samples were used directly for total DNA extraction using QIAamp UCP Pathogen Mini kit with Pathogen Lysis Tubes L (Qiagen, Hilden, Germany). Two μL extracted DNA were used per PCR primer pool for amplification and barcoding. More details on cycling conditions, library preparation and sequencing can be found in **Supplementprotocol: MultiSeq_AMR**.

### Rapid enrichment combined with MultiSe-qAMR

To accelerate reporting time, a rapid culture enrichment method was tested using eight Gram negative and seven Gram positive ATCC/clinical isolates (**SupplementRapid_enrichment_S3**). For this, between 1 and 10 CFU of log phase cells were spiked in BACT/ALERT culture bottles pre-supplemented with 10 mL sterile sheep blood. Culture bottles were incubated and taken out at every 2-hour interval for plating and colony counts for up to 10 hours using the method described by Miles & Misra.^33^ All the culture experiments were done in triplicates and during every sampling session, one mL enriched samples were aspirated from the culture bottles and preserved at −80 °C for further processing.

Finally, 13 ATCC/clinical isolates enriched for 6 hours and preserved at −80 °C were chosen to be processed further with MultiSeq-AMR. To remove unwanted host DNA that might oversaturate the PCR reaction and to concentrate bacteria in the sample, a rapid host depletion step was performed for the 6-hour enriched spiked blood (REBC) samples prior to DNA extraction (**Supplementprotocol: MultiSeq_AMR: Pre-processing REBC samples**). DNA extraction, PCR amplification and barcoding, and library preparation was performed as described in **Supplementprotocol: MultiSeq_AMR**.

### Bioinformatic analysis to identify species and AMR determinants

To classify bacterial/fungal species and AMR determinants, fastq reads with ≥Q9 scores were initially trimmed to remove barcode primers and adapters using porechop (version 0.2.4).^34^ Adapter trimmed fastq files were then processed with Kraken 2^18^ using a custom 16S rRNA, 18S rRNA, 28S rRNA and ITS database to differentiate AMR and species specific reads. Sequencing reads that matched with the Kraken 2 database were grouped as species, and unclassified reads at this stage were pooled separately as potential AMR determinants. Next, classified fastq reads were assembled *de novo* using Megahit (version 1.2.9)^35^ with default parameters, and contigs smaller than 200 bp and having a multiplicity/depth less than 20 were dropped. Next, the filtered contigs were subjected to BLASTn (version 2.15.0)^36^ using the NCBI nr database and information such as Subject Taxonomy IDs (staxids), Subject Scientific Name (ssciname), Subject Common Names (scomnames), Subject Blast Names (sblastnames), Subject Super Kingdoms (sskingdoms), Subject Sequence ID (sseqid), Bit Score (bitscore), Alignment Length (length), Expectation Value (evalue), and Percentage Identity (pident) from the top 10 hits per contig were saved for further analysis. Finally, the most common ssciname/ sscinames (representing ≥25% of all hits) per sample or barcode identified with BLASTn were reported as the causative bacterial/fungal species (**Fig. 4**).

Unclassified reads were directly analysed using ResFinder (version 4.5.0) with “fastq (Nanopore Reads) option” (“--nanopore”) to identify AMR determinants. The identity and length (-l and -t) thresholds were lowered and set to 40% to allow for identification of MultiSeq-AMR amplicons that are ≤50% of the reference gene sequences present in the ResFinder database and might be missed during the search (**Fig. 4**). Classified AMR reads were finally compared to the sequencing (WGS) results obtained from our earlier study^32^ or the ATCC/reference strains. Accuracy of MultiSeq-AMR in identifying AMR determinants was assessed using four categories: false negative, false positive, true negative, and true positive. Where, false negative means that AMR genes were targeted by the MultiSeq-AMR panel and were present in the bacteria but were not identified, false positive means that genes/targets that are absent in the reference strain but detected with MultiSeq-AMR, true negative = that are not present and not supposed to be detected, and remained undetected, true positive = genes/target that are present in both the panel and the bacteria and were identified with MultiSeq-AMR.

To understand the minimum DNA yield in Mbp required (per sample) to predict species/AMR with same/similar accuracy, all the sequencing reads were subsampled randomly (10 Mbp, 8 Mbp, 6 Mbp, 4 Mbp, and 2 Mbp) with rasusa^37^ and processed similarly. Instead using sequencing time which can vary depending on the type of sequencing flow cells used, yield in Mbp was used as more consistent proxy.

### Statistical analysis

Statistical analysis was performed, and graphs were generated using R base (version 4.3.3) in RStudio (version 2023.12.1). R packages readr, readxl, dplyr, ggplot2, stringr, reshape2, and viridis were used for this purpose.

## Data availability

Raw amplicon sequencing reads (fastq.gz) analysed and described in this study can be found on European Nucleotide Archive under the study accession “PRJEB81234”.

## Acknowledgements

We want to express our gratitude to Kareen Macleod for providing us with the ATCC reference stains tested in this study. We would also like to thank Ângelo Mendes and Ryan Carter for their continuous support and input during this project.

## Authors’ contributions

Initial concept of study was generated by MSIS and TF. Data generation, curation, formal analysis, and initial thesis draft was written by MSIS. Project investigation, experimental method selection, formal review and coordination was done by MSIS, KO, KB, PE, MEM and TF, with additional methods input and review from MJHG. Clinical sample collection and coordination was done by MEM, CW and MF.

## Ethical statement

This study was approved by UK National Health Services (NHS) Greater Glasgow and Clyde (R&I reference: GN19ID331), and the University of Glasgow College of Medical, Veterinary & Life Sciences Ethics Committee (Project No: 200210015). Blood culture samples from all the individuals suspected of having BSI were taken as a part of routine clinical care and no personal information from the patients or from the healthy human volunteers was collected and used in this study.

## Supplement protocol:MultiSeq-AMR

### Consumables

| Name | Concentration/units | Commercial source | Comments |
| --- | --- | --- | --- |
| MultiSeq primer pools | 10 µm per pool | IDT or Eurofins | Check list of primers & pools below |
| Saponin<br>(Only for REBC samples) | Final concentration 2.25% | Tokyo chemical industry, Cat: S0019 | To make a 2.25% w/v solution, dissolve 2.25 grams of Saponin into 50 ml sterile nuclease free H<sub>2</sub>O. Mix thoroughly until dissolved. Adjust the volume to 100ml. Sterilise using 0.2 µm filter, and store at 4 °C for up to 3 weeks. |
| DNase I | 300 units/µL | Invitrogen, Cat: 18047019 | NA |
| PCR H2O | NA | Qiagen, Cat: 17000-10 | NA |
| DNA binding beads | NA | Aline biosciences, Cat: C-1003-50 | NA |
| Molecular grade ethanol | NA | Sigmaaldrich, Cat: 51976-500ML-F | NA |
| Qubit dsDNA BR assay Kit | NA | ThermoFisher, Cat: Q32853 | NA |
| Qubit assay tubes | NA | ThermoFisher, Cat: Q32856 | NA |
| PCR master mix | NA | RepliQa HiFi ToughMix, Quantabio, Cat: 95200-500 | For ultra rapid PCR amplification and barcoding |
| Proteinase K | NA | Qiagen, Cat: RP107B-1 |  |
| DNA extraction kit | NA | Qiagen, Cat: 50214, with Pathogen Lysis Tubes L, Cat: 19092 | Users can choose their own extraction method as long as it is sensitive and robust |
| DNA/RNA Shield | 1x | Zymo research, Cat: R1100-50 | NA |
| Ligation Sequencing Kit | NA | ONT, Cat: SQK-LSK109 | NA |
| PCR Barcoding Expansion Kit | NA | ONT, Cat: EXP-PBC096 | NA |
| NEBNext Ultra II End Repair/dA-Tailing Module | NA | NEB, Cat: E7546L | NA |
| NEBNext Quick Ligation Module | NA | NEB, Cat: E6056L | NA |
| Flow Cell Priming Kit | NA | ONT, Cat: EXP-FLP002 | Required for both protocols |
| Flow Cell Wash Kit | NA | ONT, Cat: EXP-WSH004 | Required for both protocols |
| ONT Flow Cell R9.4.1 | NA | ONT, Cat: FLO-MIN106 or FLO-FLG001 | Required for both protocols |
| Magnetic rack/plate holder | NA | ThermoFisher, Cat: 12331D and 12321D | NA |
| PCR plate 96 well | NA | Merck, Cat: BR781378 | NA |
| 1.5 ml DNA LoBind tubes | NA | Eppendorf, Cat: 0030108051-250EA | Use LoBind tubes |
| 2 ml round bottom tubes | NA | Starlab, Cat: S1620-2700-C or Eppendorf, Cat: 0030120094 | Avoid using LoBind tubes |
| Pipettes and pipette tips P2, P10, P20, P100, P200, P1000, Multichannel | NA | Any | NA |
| Centrifuge /Microplate centrifuge | NA | Any | NA |
| Heating block for 1.5/2 mL tubes | NA | Any | NA |
| Sequencing device | NA | Flongle, MinION Mk1B, GridION | NA |
| Device for live basecalling | NA | GridION or A powerful laptop | A laptop with minimum 8 Gb GPU (RTX 3060 or higher), 16 GB Ram, 1 Tb M.2 NVMe SSD |

**Disclaimer:** Some parts of the following protocol has been copied directly from the manufacturer's official website or from the kit manual. Therefore, all the proprietary information mentioned below remains the sole property of the kit's manufacturer.

### Before sample preparation

It is important to divide all reagents into smaller aliquots to avoid multiple freeze-thaw cycles and reduce the likelihood of cross-contamination. Thaw all the reagents at room temperature or on ice as per manufacturers recommendation.

This protocol can be used on BD BACTEC (Becton, Dickinson and Company, NJ, USA) or any blood culture (FPBC) samples flagged positive with an automated culture system. Protocol M-15 mNGS can also be applied to rapid culture-enriched blood samples (REBC; for example, 1- 10 mL patient blood samples incubated for 6 hours in BD BACTEC media bottles), once the enriched samples have at least 103 CFU/mL bacterial concentration. FPBC samples can be used directly for chemical host DNA depletion (CHDD); however, REBC samples need to be pre-processed to concentrate the sample before continuing with this protocol. It is crucial to use fresh REBC samples for CHDD, as frozen or improperly stored samples can lead to the lysis of bacterial cells. This may lead to a considerable loss of bacterial DNA during CHDD.

### Pre-processing REBC samples

1. Transfer 1.5 mL REBC samples carefully to a 2 mL round bottom microcentrifuge tube and centrifuge at 8,000 xg for 5 minutes. Note* It is important to use round/wide bottom 2 mL microcentrifuge tubes to minimise bacterial cell loss while aspirating the supernatant during the initial washing steps.
2. Slowly aspirate the supernatant and resuspend the pellet in 800 μL 2.25% saponin solution.
3. Add 1 μL DNase I (300 units/μL) to the sample and incubate at 25°C in a shaking heating block for 10 minutes at 800 rpm.
4. Centrifuge the tube at 10,000 xg for 5 minutes, aspirate the supernatant carefully and resuspend the pellet in 1x DNA/RNA Shield according to the input volume for the DNA extraction kit (for example, 400 μL for QIAamp UCP kit).

### DNA extraction (Mechanical Pr-elysis Protocol for whole blood)

400 μL FPBC or REBC samples in 1x DNA/RNA Shield can be extracted using QIAamp UCP Pathogen Mini kit with Pathogen Lysis Tubes L following manufacturer’s instructions. Following extraction and purification steps, elute the DNA in 50 μL Buffer AVE and then proceed to the rapid PCR barcoding step.

Note* Alternative bacterial cell lysis and DNA extraction methods (e.g., automated or faster) can also be used, however, for the rapid enrichment method, it is important that the users test the efficiency in lysing tough bacterial cells and DNA recovery rate/analytical sensitivity for low abundant samples before proceeding.

Protocol for mechanical pre lysis (Pathogen Lysis Tubes L) with QIAamp UCP Pathogen Mini kit can be accessed from the link below.

https://www.qiagen.com/us/resources/download.aspx?id=0930379e-a52a-475a-acc5-d944701dbaeb&lang=en

### Rapid PCR barcoding of AMR and specie-specific targets

Combine 100 µM stock primers (forward and reverse) in equal volumes (e.g., 10 µL each) for each primer according to the following combinations for each pool:

| Pool 1 | Pool 2 | Pool 3 | Pool 4 | Pool 5 | Pool 6 | Pool 7 | 16S + 28S |
| --- | --- | --- | --- | --- | --- | --- | --- |
| *blaNDM* | *blaSHV* | *blaTEM* | *blaPER* | *cmlA* | *ErmA* | *dfrA1* | 16S rRNA |
| *blaKPC* | *MCR-1* | *blaOXA-9* | *blaCMY1/MOX* | *sul1* | *ErmB* | *dfrA12* | NL1-NL4 |
| *blaIMP* | *MCR-2* | *blaOXA-23* | *blaCMY-2* | *sul2* | *aph(3')-III* | *dfrA7/17* | NA |
| *blaVIM* | *VanA* | *blaOXA-24* | *AAC(6')-Ib* | *ant(2'')-I* | *aph(3')-IIa* | *Str* | NA |
| *blaGES* | *VanB* | *blaOXA-58* | *qnrB* | *aac(3)-II* | *aph(3')-Ia* | *catA1* | NA |
| *blaOXA-48* | *MecA* | *blaFOX* | *qnrS* | *aac(3)-IV* | *aadA1* | *catB7* | NA |
| *blaCTX-Group-M-All* | *MecC* | *blaACC* | *qepA* | *mphA* | *aadA2* | *oqxA* | NA |
| Pool 1 to Pool 7 and 16S+28S tested in this study |  |  |  |  |  |  |  |

| Pool 8 | Pool 9 | Pool 10 | Pool 11 | Pool 12 | Pool 13 |
| --- | --- | --- | --- | --- | --- |
| *blaOXA-1* | *blaGIM* | *gyrA* | *msrA* | *parC* | *tet(A)* |
| *blaOXA-7* | *blaSPM* | *ermC* | *msrC* | *oqxB* | *tet(B)* |
| *blaOXA-50* | *blaVEB* | *ermF* | *msrD* | *catA2* | *tet(C)* |
| *blaOXA-51* | *blaBEL* | *ermG* | *msrE* | *sul3* | *blaDHA* |
| *qnrA* | *mcr-3* | *ermT* | *cmlB* | *mefA* | *blaACT/MIR* |
| *qnrC* | *mcr-4* | *mphB* | *rmtB* | *mefB* | *aph(3')-Ib* |
| *qnrD* | *mcr-5* | *mphE* | *armA* | *mefE* | *dfrA5* |
| Pool 8 to Pool 14 not tested in this study |  |  |  |  |  |

Dilute the 100 µM pools at 1:10 ratio in molecular grade nuclease free water to create stocks of 10 µM primer pools. Primers are used at a final concentration of 50 nanomolar (nM) per primer in MultiSeq-AMR protocol. AMR primers are combined to target up to 7 AMR targets per pool (can be extended further). The primer targets and/or pools are modular and can be customised/expanded (from 8 pools, and 12 samples to 12 pools, 8 samples per PCR plate) according to requirements.

1. Combine the following components using a multichannel pipette and dispense into a 0.2 mL PCR plate. For example, for Sample1 using Barcode1: (Up to 96 samples, can be processed per batch). Note* As an alternative, liquid handlers (conventional or acoustic) can be used for 0.5x, 0.25x volume and for faster aliquoting.

| Reagent (μL) 1x | Pool 1 | Pool 2 | Pool 3 | Pool 4 | Pool 5 | Pool 6 | Pool 7 | 16S + 28S |
| --- | --- | --- | --- | --- | --- | --- | --- | --- |
| RepliQa HiFi ToughMix (2x) | 10 | 10 | 10 | 10 | 10 | 10 | 10 | 10 |
| MultiSeq primers (10 μM) | 1.4 | 1.4 | 1.4 | 1.4 | 1.4 | 1.4 | 1.4 | 0.4 |
| PCR barcodes (10 μM) | 1 | 1 | 1 | 1 | 1 | 1 | 1 | 1 |
| Purified DNA | 3 | 3 | 3 | 3 | 3 | 3 | 3 | 3 |
| PCR H<sub>2</sub>O | 4.6 | 4.6 | 4.6 | 4.6 | 4.6 | 4.6 | 4.6 | 5.6 |
| Total | 20 | 20 | 20 | 20 | 20 | 20 | 20 | 20 |

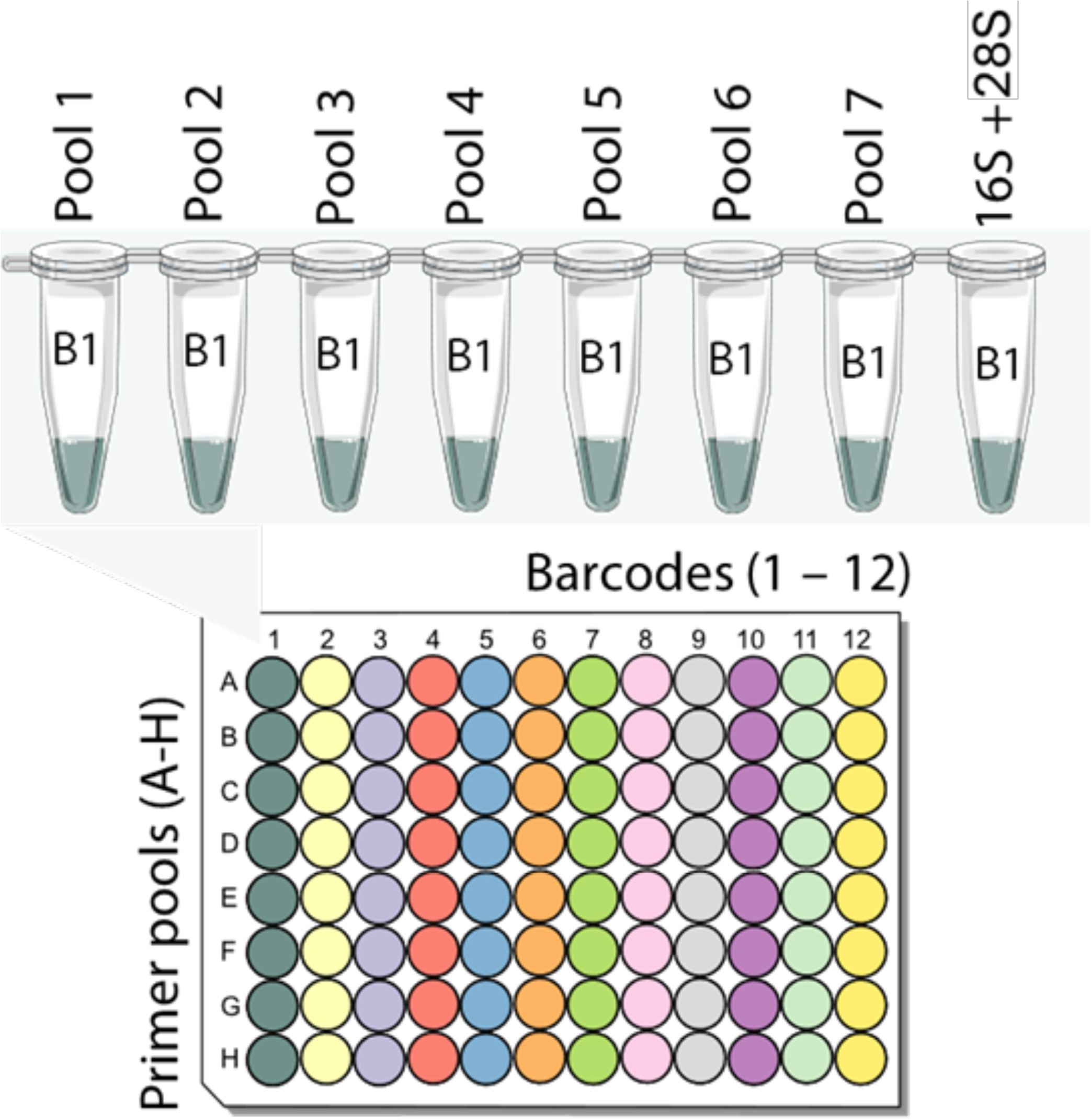
2. Seal the plate tightly, spin and perform PCR with the following conditions with the heated lid on.

3. Following PCR, spin down the materials by performing a quick spin and combine pools 1 through 8 using a multichannel pipette. Take 5 μL PCR product from pools 1 to 7 and 10 μL (half) from the 16S/28S pool.
4. Resuspend the DNA binding beads (e.g., AMPure XP or Aline biosciences) by vortexing. Note* DNA binding beads can be aliquoted in PCR strips. This can be done upon receiving all the reagents, so these steps do not take addition time when processing the samples.
5. Add 2 µL of 20 mg/mL proteinase K to each sample tube using a multichannel pipette. Note* For this 20 mg/mL proteinase K needs to be aliquoted in PCR strips. This can be done upon receiving all the reagents, so these steps do not take addition time when processing the samples.

| Step | Primers active | Temp (°C) | Time (seconds) | Cycles |
| --- | --- | --- | --- | --- |
| Initial denaturation | None | 98 | 120 | 1 |
| Denaturation | AMR/species | 98 | 10 | 5-7* |
| Annealing |  | 58 | 10 |  |
| Extension |  | 68 | 15 |  |
| Denaturation | AMR/species | 98 | 10 | 5-7* |
| Annealing |  | 57 | 10 |  |
| Extension |  | 68 | 15 |  |
| Denaturation | AMR/species | 98 | 10 | 5-7* |
| Annealing |  | 56 | 10 |  |
| Extension |  | 68 | 15 |  |
| Denaturation | AMR/species | 98 | 10 | 5-7* |
| Annealing |  | 55 | 10 |  |
| Extension |  | 68 | 15 |  |
| Denaturation | Barcode | 98 | 10 | 10-12* |
| Annealing |  | 62 | 10 |  |
| Extension |  | 68 | 15 |  |
| Final extension | None | 68 | 60 | 1 |

Note\* For FPBC samples, perform (5x4 = 20 + 10) a total of 30 PCR cycles. For REBC, perform (7x4 = 28 + 12) 40 PCR cycles overall.

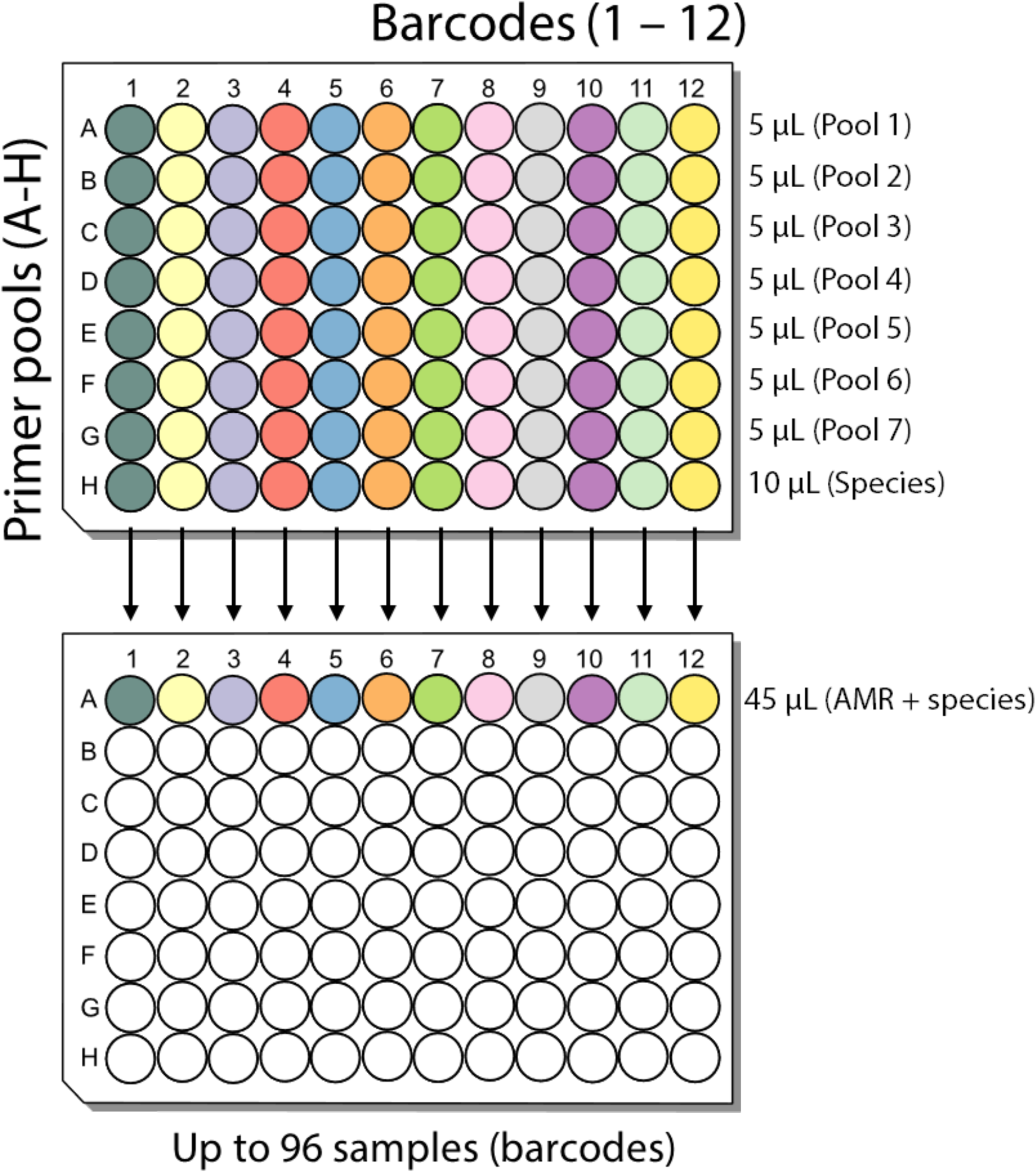
6. Similarly, add 23 µL bead suspension (approximately 0.5x) per sample using a multichannel pipette and mix by pipetting up and down.
7. Incubate the sample for 5 minutes at room temperature.
8. Pellet the sample on a magnetic stand until the supernatant is clear and colourless. Pipette off the supernatant by keeping the tube on the magnet.
9. Keep the plate on the magnetic stand and wash the beads with 200 µL of freshly prepared 70% ethanol without disturbing the pellet. Aspirate the ethanol carefully without touching the pellet using a multichannel pipette and discard.
10. Repeat the previous ethanol washing step.
11. Pipette off any residual ethanol by keeping the tube on the magnet. Allow to air dry for about ∼30 seconds, but do not dry the pellet to the point of cracking.
12. Remove the plate from the magnetic stand and resuspend pellet in 25 µL nuclease-free water. Incubate for 2 min at room temperature.
13. Pellet the beads on a magnet until the eluate is clear and colourless (at least for one minute).
14. Remove and retain 20 µL of eluate into a clean PCR plate.
15. Quantify 1 µL barcoded DNA from each sample using a Qubit fluorometer.

### Ligation sequencing

#### End prep

1. Pool up to 96 samples in a 0.2 mL PCR tube, ensuring the total DNA amount is exactly 200 ng (or approximately 400 fmol, assuming a fragment size of 0.75 kb) in a volume of 49 µl, using the following procedure. The example below is showing pooling procedure for 12 samples.

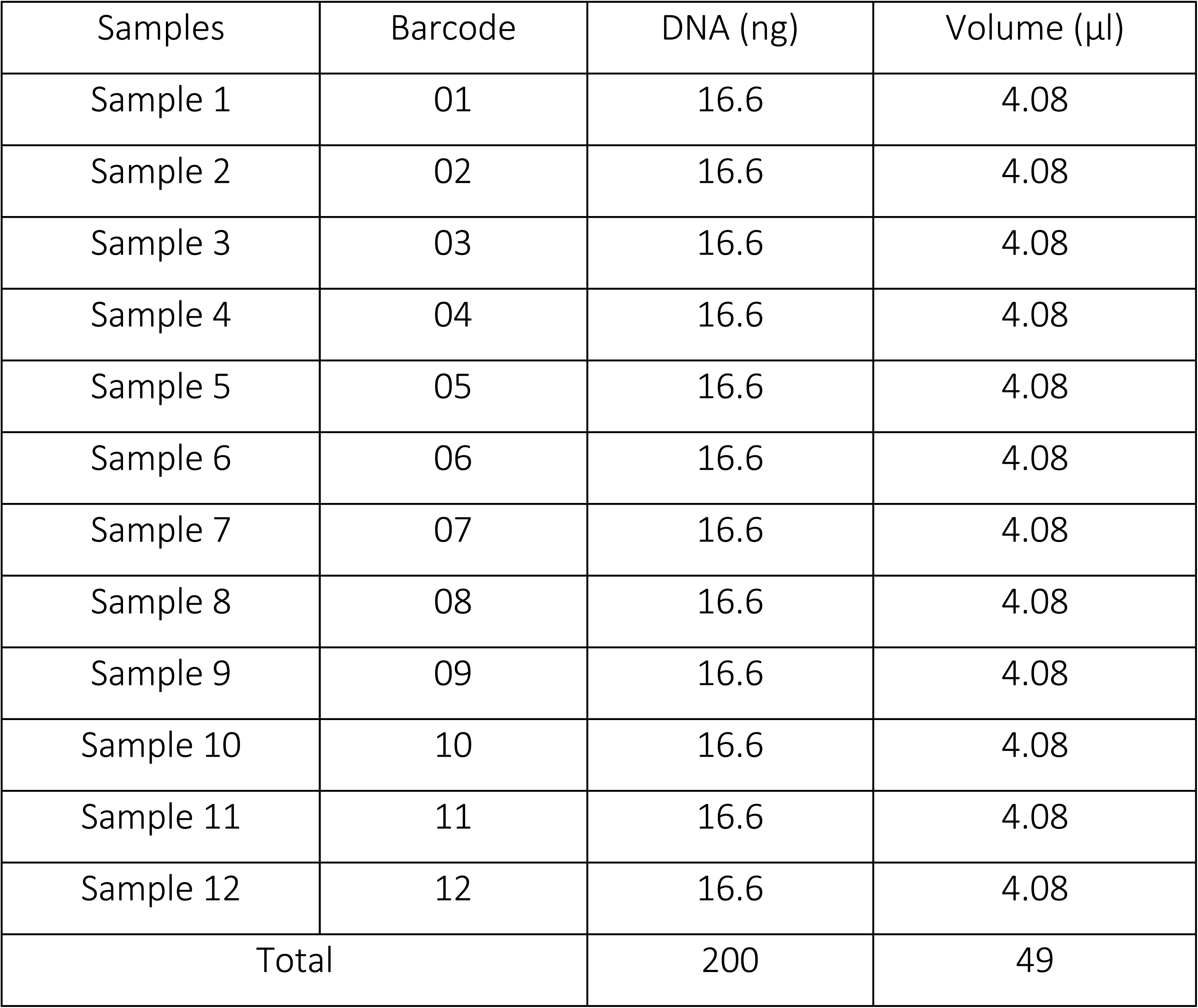
2. Now combine the following reagents in a 0.2 mL PCR tube.

3. Mix the samples gently by flicking the tube multiple times, and spin down.
4. Incubate the sample at 20°C in a thermal cycler for 5 minutes with heated lid option turned off.
5. Next, incubate sample again at 65°C for 5 minutes with heated lid on. Native barcode ligation and cleanup
6. Combine the following reagents in a 0.2 mL PCR tube.

2. Now combine the following reagents in a 0.2 mL PCR tube.
| Reagent | Volume (µL) |
| --- | --- |
| DNA control sample (DCS) | 1 |
| Ultra II End-prep Reaction Buffer | 7 |
| Ultra II End-prep Enzyme Mix | 3 |
| Pooled DNA (200 ng) | 49 |
| Total | 60 |

7. Flick to mix the sample multiple times, spin and incubate the reaction mixture for 10 minutes at 20 °C.
8. Resuspend the DNA binding beads (e.g., AMPure XP or Aline biosciences) by vortexing.
9. Add 100 µL adapter ligated DNA sample to 50 µL (0.5x) of resuspended DNA binding beads in a 1.5 mL Eppendorf DNA LoBind tube and mix by pipetting.
10. Incubate the mixture for 5 minutes at room temperature.
11. Spin down the sample and pellet on a magnetic rack until the supernatant is clear and colourless. Keep the tube on the magnet, and carefully pipette off the supernatant.
12. Wash the beads by adding 250 μl Short Fragment Buffer (SFB).
13. Repeat the previous SFB washing step.
14. Spin down and place the tube back on the magnetic rack. Pipette off any residual SFB. Allow the tube to dry for ∼30 seconds, but do not dry the pellet to the point of cracking.
15. Remove the tube from the magnetic rack and resuspend the pellet in 17 µL Elution Buffer (EB).
16. Spin down briefly and incubate the mixture for 2 minutes at room temperature.
17. Pellet the beads on a magnetic rack until the eluate is clear and colourless, for at least 1 minute.
18. Remove and retain 15 µL of the eluate (EB) containing the DNA library into a clean 1.5 mL Eppendorf DNA LoBind tube.
19. For MinION flow cell, load approximately 25 ng (51.36 fmol; considering 0.75kb) of final prepared library onto the MinION flow cell following manufacturers recommendation. Alternatively, if processing a few samples per batch, load 10 ng (20.54 fmol; considering 0.75kb) of final prepared library onto the Flongle flow cell.

| Reagent | Volume (μL) |
| --- | --- |
| End prepped DNA | 60 |
| Adapter Mix (AMX or AMX-F) | 5 |
| Ligation Buffer (LNB) | 25 |
| Quick T4 DNA Ligase | 10 |
| Total | 100 |

Note* The typical library size after this stage is around 0.75 kb. However, users can verify this by running the library on a gel or TapeStation for a few batches. Once the average expected library size is determined (sample or site-wise), there will be no need to repeat this for every batch of samples.

For loading sequencing library or washing flow cells, follow the manufacturers recommended protocols.

### Priming and loading sequencing libraries

https://nanoporetech.com/document/rapid-barcoding-sequencing-sqk-rbk004#priming-and-loading-the-sp

https://community.nanoporetech.com/nanopore_learning/lessons/priming-and-loading-your-flow-cell

### Washing flow cells for re-use

https://nanoporetech.com/document/flow-cell-wash-kit-exp-wsh004

## References

1. Diekema, D.J. et al. The Microbiology of Bloodstream Infection: 20-Year Trends from the SENTRY Antimicrobial Surveillance Program. Antimicrob Agents Chemother 63, 10.1128/aac.00355-19 (2019).

2. Ikuta, K.S., Lucien R. Swetschinski, Gisela R. Aguilar, Fablina Sharara, Tomislav Mestrovic, Authia P. Gray, Nicole D. Weaver. Global mortality associated with 33 bacterial pathogens in 2019: a systematic analysis for the Global Burden of Disease Study 2019. Lancet 400, 2221–2248 (2022).

3. Sakr, Y. et al. Sepsis in Intensive Care Unit Patients: Worldwide Data From the Intensive Care over Nations Audit. Open Forum Infect Dis 5, ofy313 (2018).

4. Scheer, C.S. et al. Impact of antibiotic administration on blood culture positivity at the beginning of sepsis: a prospective clinical cohort study. Clin Microbiol Infect 25, 326–331 (2019).

5. Strich, J.R., Heil, E.L. & Masur, H. Considerations for Empiric Antimicrobial Therapy in Sepsis and Septic Shock in an Era of Antimicrobial Resistance. J Infect Dis 222, S119–S131 (2020).

6. Ohnuma, T. et al. Association of Appropriate Empirical Antimicrobial Therapy With In- Hospital Mortality in Patients With Bloodstream Infections in the US. JAMA Netw Open 6, e2249353 (2023).

7. Salam, M.A. et al. Antimicrobial Resistance: A Growing Serious Threat for Global Public Health. Healthcare (Basel) 11, 19–46 (2023).

8. Paharik, A.E., Schreiber, H.L.t., Spaulding, C.N., Dodson, K.W. & Hultgren, S.J. Narrowing the spectrum: the new frontier of precision antimicrobials. Genome Med 9, 110 (2017).

9. D’Andreano, S., Cusco, A. & Francino, O. Rapid and real-time identification of fungi up to species level with long amplicon nanopore sequencing from clinical samples. Biol Methods Protoc 6, bpaa026 (2021).

10. Ali, J., Johansen, W. & Ahmad, R. Short turnaround time of seven to nine hours from sample collection until informed decision for sepsis treatment using nanopore sequencing. Sci Rep 14, 6534 (2024).

11. Bauer, M.J. et al. Optimized Method for Bacterial Nucleic Acid Extraction from Positive Blood Culture Broth for Whole-Genome Sequencing, Resistance Phenotype Prediction, and Downstream Molecular Applications. J Clin Microbiol 60, e0101222 (2022).

12. Sajib, M.S.I. et al. Advances in host depletion and pathogen enrichment methods for rapid sequencing-based diagnosis of bloodstream infection. The Journal of Molecular Diagnostics 9, 741–753 (2024).

13. Zhang, Y., Lu, X., Tang, L.V., Xia, L. & Hu, Y. Nanopore-Targeted Sequencing Improves the Diagnosis and Treatment of Patients with Serious Infections. mBio 14, e0305522 (2023).

14. Zhao, K. et al. Rapid Identification of Drug-Resistant Tuberculosis Genes Using Direct PCR Amplification and Oxford Nanopore Technology Sequencing. Can J Infect Dis Med Microbiol 2022 (**1**), 7588033 (2022).

15. Zhang, C. et al. Multiplex PCR and Nanopore Sequencing of Genes Associated with Antimicrobial Resistance in Neisseria gonorrhoeae Directly from Clinical Samples. Clin Chem 67, 610–620 (2021).

16. Smith, S.D. et al. Diversity of Antibiotic Resistance genes and Transfer Elements-Quantitative Monitoring (DARTE-QM): a method for detection of antimicrobial resistance in environmental samples. Commun Biol 5, 216 (2022).

17. Li, Y. et al. Multiplexed Target Enrichment Enables Ejicient and In-Depth Analysis of Antimicrobial Resistome in Metagenomes. Microbiol Spectr 10, e0229722 (2022).

18. Wood, D.E., Lu, J. & Langmead, B. Improved metagenomic analysis with Kraken 2. Genome Biol 20, 257 (2019).

19. Li, H. Minimap2: pairwise alignment for nucleotide sequences. Bioinformatics 34, 3094–3100 (2018).

20. Somily, A.M. et al. Time-to-detection of bacteria and yeast with the BACTEC FX versus BacT/Alert Virtuo blood culture systems. Ann Saudi Med 38, 194–199 (2018).

21. Alcock, B.P. et al. CARD 2023: expanded curation, support for machine learning, and resistome prediction at the Comprehensive Antibiotic Resistance Database. Nucleic Acids Res 51, D690–D699 (2023).

22. Bortolaia, V. et al. ResFinder 4.0 for predictions of phenotypes from genotypes. J Antimicrob Chemother 75, 3491–3500 (2020).

23. Clausen, P., Aarestrup, F.M. & Lund, O. Rapid and precise alignment of raw reads against redundant databases with KMA. BMC Bioinformatics 19, 307 (2018).

24. Callahan, B.J. et al. High-throughput amplicon sequencing of the full-length 16S rRNA gene with single-nucleotide resolution. Nucleic Acids Res 47, e103 (2019).

25. Romanelli, A.M., Fu, J., Herrera, M.L. & Wickes, B.L. A universal DNA extraction and PCR amplification method for fungal rDNA sequence-based identification. Mycoses 57, 612–22 (2014).

26. Edgar, R.C. MUSCLE: multiple sequence alignment with high accuracy and high throughput. Nucleic Acids Res 32, 1792–7 (2004).

27. Ye, J. et al. Primer-BLAST: a tool to design target-specific primers for polymerase chain reaction. BMC Bioinformatics 13, 134 (2012).

28. Boyd, D.A. et al. Complete nucleotide sequence of a 92-kilobase plasmid harboring the CTX-M-15 extended-spectrum beta-lactamase involved in an outbreak in long-term-care facilities in Toronto, Canada. Antimicrob Agents Chemother 48, 3758–64 (2004).

29. Jensen, L.B., Frimodt-Moller, N. & Aarestrup, F.M. Presence of erm gene classes in gram-positive bacteria of animal and human origin in Denmark. FEMS Microbiol Lett 170, 151–8 (1999).

30. Bansal, S. & Tandon, V. Contribution of mutations in DNA gyrase and topoisomerase IV genes to ciprofloxacin resistance in Escherichia coli clinical isolates. Int J Antimicrob Agents 37, 253–5 (2011).

31. Ng, L.K., Martin, I., Alfa, M. & Mulvey, M. Multiplex PCR for the detection of tetracycline resistant genes. Mol Cell Probes 15, 209–15 (2001).

32. Sajib, M.S.I. et al. Rapid and modular workflows for same-day sequencing-based detection of bloodstream infections and antimicrobial resistance determinants. medRxiv, 2024.10. 09.24315014 (2024).

33. Miles, A.A., Misra, S.S. & Irwin, J.O. The estimation of the bactericidal power of the blood. J Hyg (Lond) 38, 732–49 (1938).

34. Wick, R.R., Judd, L.M., Gorrie, C.L. & Holt, K.E. Completing bacterial genome assemblies with multiplex MinION sequencing. Microb Genom 3, e000132 (2017).

35. Li, D., Liu, C.M., Luo, R., Sadakane, K. & Lam, T.W. MEGAHIT: an ultra-fast single-node solution for large and complex metagenomics assembly via succinct de Bruijn graph. Bioinformatics 31, 1674–6 (2015).

36. Altschul, S.F., Gish, W., Miller, W., Myers, E.W. & Lipman, D.J. Basic local alignment search tool. J Mol Biol 215, 403–10 (1990).

37. Hall, M. Rasusa: Randomly subsample sequencing reads to a specified coverage. Journal of Open Source Software 7, 3941 (2022).

